# Synaptic loss in primary tauopathies revealed by [_11_C]UCB-J positron emission tomography

**DOI:** 10.1101/2020.01.24.20018697

**Authors:** Negin Holland, P. Simon Jones, George Savulich, Julie K. Wiggins, Young T. Hong, Tim D. Fryer, Roido Manavaki, Selena Milicevic Sephton, Istvan Boros, Maura Malpetti, Frank H. Hezemans, Franklin I. Aigbirhio, Jonathan P. Coles, John O’Brien, James B. Rowe

## Abstract

**Background:** Synaptic loss is a prominent and early feature of many neurodegenerative diseases.

**Objectives:** We tested the hypothesis that synaptic density is reduced in the primary tauopathies of progressive supranuclear palsy (PSP-Richardson’s syndrome) and amyloid-negative corticobasal syndrome (CBS).

**Methods:** Forty four participants (15 CBS, 14 PSP, and 15 age-/sex-/education-matched controls) underwent positron emission tomography (PET) with the radioligand [_11_C]UCB-J, which binds to synaptic vesicle glycoprotein 2A (SV2A), a marker of synaptic density; participants also had 3T magnetic resonance imaging and clinical and neuropsychological assessment.

**Results:** Nine CBS patients had negative amyloid biomarkers determined by [_11_C]PiB PET and hence were deemed likely to have corticobasal degeneration (CBD). Patients with PSP-Richardson’s syndrome and amyloid-negative CBS were impaired in executive, memory and visuospatial tasks. [_11_C]UCB-J binding was reduced across frontal, temporal, parietal, and occipital lobes, cingulate, hippocampus, insula, amygdala and subcortical structures in both PSP and CBD patients compared to controls (p<0.01), with median reductions up to 50%, consistent with post mortem data. Reductions of 20-30% were widespread even in the areas of the brain with minimal atrophy. There was a negative correlation between global [_11_C]UCB-J binding and the PSP and CBD rating scales (R= −0.61 p<0.002, R= −0.72 p<0.001, respectively) and a positive correlation with the revised Addenbrookes Cognitive Examination (R=0.52, p=0.01).

**Conclusions:** We confirm severe synaptic loss in PSP and CBD in proportion to disease severity, providing critical insight into the pathophysiology of primary degenerative tauopathies. [_11_C]UCB-J may facilitate treatment strategies for disease-modification, synaptic maintenance or restoration.

## Introduction

The primary degenerative tauopathies of Progressive Supranuclear Palsy (PSP) and Corticobasal Degeneration (CBD) cause a severe combination of movement and cognitive impairment (1-4). Pathologically both are associated with a 4-repeat tauopathy (5). We proposed that the neurophysiological and functional impairments in PSP and CBD are at least in part a consequence of synaptic loss. For example, at *post mortem* there is approximately 50% loss of cortical synapses in PSP and CBD (6,7), and *in vivo* there is limited evidence of a ~20% loss of post-synaptic GABAa receptors as shown with [_11_C]flumazenil positron emission tomography (PET) (8,9). Indeed abnormal physiology in pathways involved in presynaptic function have been identified from transcriptomic studies in patients with mutations in the microtubule-associated protein tau (MAPT) gene (10). Transgenic models of tauopathies (e.g. rTg4510) confirm a synaptotoxic effect of oligomeric tau, before cell death (11,12). Moreover, in other neurodegenerative dementias, such as Alzheimer’s disease, synaptic loss correlates better with cognitive dysfunction than atrophy (13).

We therefore tested the hypothesis that PSP and CBD reduce synaptic density, in proportion to disease severity. We include patients with the classic phenotype of PSP, PSP-Richardson’s syndrome, which has a high clinicopathological correlation (14), and presents with postural instability, supranuclear gaze palsy, axial rigidity and cognitive impairment (15). Other phenotypes of PSP are increasingly recognised (3,16), but excluded here. We include patients with Corticobasal Syndrome (CBS), with combinations of asymmetric rigidity, apraxia, dystonia, alien limb, and cognitive impairment (1,17). In order to identify those with probable underlying CBD, it is necessary to exclude the substantial minority of CBS caused by Alzheimer’s disease pathology (18). We therefore used amyloid imaging to distinguish those with CBS due to CBD, *versus* Alzheimer’s disease; we refer to this group as the CBD cohort. Both PSP and CBD are associated with cortical and subcortical atrophy on magnetic resonance imaging (MRI) (19); and changes in neurophysiology and connectivity measured by magnetoencephalography and functional MRI (20-23). However, functional changes are also seen in the areas of the brain that are minimally atrophic.

We used PET with the radioligand [_11_C]UCB-J ((*R*)-1-((3-(methyl-nC)pyridin-4-yl)methyl)-4-(3,4,5-trifluorophenyl)pyr-rolidin-2-one) (24). This ligand quantifies synaptic density (25,26) based on its affinity for the presynaptic vesicle glycoprotein 2A (SV2A), that is ubiquitously expressed in all brain synapses (27,28). [_11_C]UCB-J has revealed hippocampal synaptic loss in Alzheimer’s disease, correlating with episodic memory loss and clinical dementia severity (29). We sought correlations between regional [_11_C]UCB-J binding potentials, a metric of synaptic density, and disease severity, in terms of cognitive decline and global impairment on the PSP and CBD rating scales.

## Methods

### Participants & Study Design

Fourteen patients with PSP-Richardson’s syndrome and fifteen patients with CBS were recruited from a tertiary specialist clinic for PSP/CBS at the Cambridge University Centre for Parkinson-Plus. Fifteen healthy volunteers were recruited from the UK National Institute for Health Research Join Dementia Research (JDR) register. Patients had either probable PSP–Richardson Syndrome (3), or both probable CBS and probable CBD (1). Healthy controls and patient volunteers were initially screened by telephone; our exclusion criteria were: current or recent history (within the last 5 years) of cancer, concurrent use of the medication levetiracetam, history of ischaemic or haemorrhagic stroke evident on MRI available from the clinic, any severe physical illness or co-morbidity that limited ability to fully participate in the study, and any contraindications to performing MRI. Eligible participants were invited for a research visit where they underwent clinical and cognitive assessment including measures of disease severity (Table 1); these included a neurological examination by a clinician including the PSP and CBD rating scales, the Unified Parkinson’s Disease Rating scale (motor subsection III), the Schwab and England Activities of Daily Living (SEADL) and Clinical Dementia Rating Scale (CDR); cognitive testing included the revised Addenbrooke’s Cognitive Examination (ACE-R), the Mini-mental State Examination (MMSE), the Montreal Cognitive Assessment (MoCA), and the INECO frontal assessment test. Patients’ carers completed the revised Cambridge Behavioural Inventory (CBI).

**Table 1.** Demographics and neuropsychological profile for each participant cohort

|  | **Control** | **CBD** | **PSP** | **F (*p*)** |
| --- | --- | --- | --- | --- |
| **M:F** | 7:8 | 7:2 | 7:7 | ns<sup>a</sup> |
| **Age at [<sup>11</sup>C]UCB-J PET in years** | 68 (7.45) | 70.56 (8.23) | 72.79 (7.74) | ns |
| **Disease duration in years** | NA | 3.94 (2.2) | 4.28 (2.57) | ns<sup>b</sup> |
| **Education in years** | 13.69 (2.66) | 12.78 (3.27) | 12.77 (5.43) | ns |
| **ACE-R total (max. 100)** | 96.47 (2.88) | 81.56 (10.83) | 80.57 (15.02) | 9.61 (<.0004) |
| Attention_Orientation (max .18) | 17.87 (0.35) | 16.89 (1.05) | 16.43 (2.06) | 4.11 (0.02) |
| Memory (max .26) | 24.53 (1.85) | 20.67 (5.66) | 21.43 (4.27) | 3.50 (0.04) |
| Fluency (max .14) | 12.80 (1.15) | 8.22 (2.86) | 6.43 (3.44) | 22.81 (<.001) |
| Language (max .26) | 25.53 (0.92) | 22.44 (5.34) | 23.43 (5.21) | ns |
| Visuospatial (max .16) | 15.73 (0.59) | 13.33 (2.55) | 12.86 (3.98) | 4.46 (0.02) |
| **MMSE (max. 30)** | 29.27 (1.33) | 26.44 (3.13) | 27.00 (2.88) | 4.78 (0.01) |
| **UPDRS (max. 132)** | 0 (0) | 38.22 (14.81) | 32.36 (16.38) | ns<sup>b</sup> |
| **PSPRS (max. 100)** | 0.13 (0.52) | 26.78 (9.61) | 29.21 (10.27) | ns<sup>b</sup> |
| **CBDRS (max. 124)** | 0.20 (0.77) | 29.12 (13.52) | 36.80 (20.41) | ns<sup>b</sup> |
| **MoCA (max. 30)** | 27.80 (1.74) | 12.25 (12.96) | 22.46 (2.96) | 8.39 (<.001) |
| **Ineco (max. 30)** | 26.00 (1.85) | 14.60 (8.47) | 17.70 (4.74) | 17.22 (<.001) |
| **CDR sum of boxes (max. 32)** | 0.07 (0.26) | 6.78 (4.71) | 7.54 (6.55) | 11.28 (<.001) |
| **CBI (max. 180)** | 2.47 (4.81) | 27.44 (13.5) | 42.43 (38.13) | 9.97 (<.001) |
| **SEADL (max. 1)** | 0.99 (0.03) | 0.56 (0.28) | 0.60 (0.22) | 10.54 (<.001) |

The results are given as mean (standard deviation). CBD here refers to corticobasal syndrome with a negative amyloid biomarker from [<sup>11</sup>C]PiB PET, and PSP refers to patients with PSP-Richardson's syndrome. The F-statistic and p-values are derived from ANOVA. ACE-R: revised Addenbrooke's Cognitive Examination, MMSE: Mini-mental State Examination, UPDRS: Unified Parkinson's Disease Rating Scale, PSPRS: Progressive Supranuclear Palsy Rating Scale, CBDRS: CBD functional rating scale, MoCA: Montreal Cognitive Assessment, Ineco: frontal assessment tool, CDR: Clinical Dementia Rating Scale, CBI: revised Cambridge Behavioural Inventory, SEADL: Schwab and England Activities of Daily Living Scale. <sup>a</sup> chi-
squared test. <sup>b</sup> ANOVA with PSP and CBD patients only. ns = non-significant at $p < 0.05$ ; NA = non-applicable.

All participants underwent simultaneous 3T MRI and [_11_C]UCB-J PET. Patients with CBS also underwent amyloid PET imaging using Pittsburgh Compound B ([_11_C]PiB) and cortical standardised uptake value ratio (SUVR; 50-70 minutes post injection; whole cerebellum reference tissue) was determined using the Centiloid Project methodology (30). Only those with a negative amyloid status as characterised by a cortical [_11_C]PiB SUVR less than 1.21 (obtained by converting the Centiloid cut-off of 19 to SUVR using the Centiloid-to-SUVR transformation) (31), are included in the subsequent analysis, with the aim of excluding patients with CBS due to Alzheimer’s disease. We interpret this amyloid-negative group as having CBD, although acknowledge that other pathologies are possible.

The research protocol was approved by the local Cambridge Research Ethics Committee (REC: 18/EE/0059) and the Administration of Radioactive Substances Advisory Committee. All participants provided written informed consent in accordance with the Declaration of Helsinki.

### Neuroimaging

[_11_C]UCB-J was synthesised at the Radiopharmacy Unit, Wolfson Brain Imaging Centre, Cambridge University, using the methodology previously described (32). Dynamic PET data acquisition was performed on a GE SIGNA PET/MR (GE Healthcare, Waukesha, USA) for 90 minutes starting immediately after [_11_C]UCB-J injection (median injected activity: 351 ± 107 MBq, injected mass ≤ 10 μg), with attenuation correction including the use of a multi-subject atlas method (33,34) and also improvements to the MRI brain coil component (35). Each emission image series was aligned using SPM12 (www.fil.ion.ucl.ac.uk/spm/software/spm12/) then rigidly registered to a T1-weighted MRI acquired during PET data acquisition (TR = 3.6 msec, TE = 9.2 msec, 192 sagittal slices, in plane resolution 0.55 × 0.55 mm (subsequently interpolated to 1.0 × 1.0 mm); slice thickness 1.0 mm). Using a version of the Hammersmith atlas (http://brain-development.org) with modified posterior fossa regions, combined regions of interest (including aggregated regions for frontal, parietal, occipital, and temporal lobes; cingulate; and cerebellum) were spatially normalized to the T1-weighted MRI of each participant using Advanced Normalisation Tools (ANTs) software (36). Regional time-activity curves were extracted following the application of geometric transfer matrix (GTM) partial volume correction (PVC, (37)) to each of the dynamic PET images. Regions of interest (ROIs) were multiplied by a binary grey matter mask (>50% on the SPM12 grey matter probability map smoothed to PET spatial resolution), with the exception of the pallidum, substantia nigra, pons and medulla because masking eliminated the ROI for some or all of the subjects. Multiple background grey matter, white matter and cerebrospinal fluid (CSF) regions were also defined to provide whole brain coverage for GTM PVC. The mean grey matter/(grey matter white matter) fraction in the masked ROIs was 0.97 ± 0.03, 0.96 ± 0.03 and 0.96 ± 0.03 for the control, CBD and PSP groups, respectively, illustrating the predominance of grey matter in the masked ROIs. To assess the impact of PVC, time-activity curves were also extracted from the same ROIs without the application of GTM PVC.

To quantify SV2A density, [_11_C]UCB-J non-displaceable binding potential (BP_ND_) was determined, both regionally and at the voxel level, using a basis function implementation of the simplified reference tissue model (38), with the reference tissue defined in the centrum semiovale (39,40). The volume-weighted average of the GTM PVC BP_ND_ values in the masked ROIs was used as a global BP_ND_ metric. Group average BP_ND_ images (illustrated in Figure 1A) were obtained by spatially normalising each individual T1-weighted MRI (and thereby the co-registered BP_ND_ map) to MNI space, and then to the group template using ANTs.

### Statistical analysis

Statistical analyses used R (version 3.6.2), with analysis of covariance (ANCOVA) to compare regional [_11_C]UCB-J BP_ND_ between the three groups (Control, CBD, PSP), with age as a covariates of no interest. Regions of interest were: frontal, temporal, parietal, occipital lobes; cingulate cortex, hippocampus, insula, amygdala, caudate nucleus, nucleus accumbens, putamen, pallidum, thalamus, cerebellum, substantia nigra, midbrain, pons, and medulla.

The relationships between [_11_C]UCB-J BP_ND_, disease severity (PSP and CBD Rating Scales) and cognition (revised Addenbrooke’s Cognitive Examination) were tested through linear models of the patient data, with age as a covariate of no interest.

The primary analyses used BP_ND_ determined following GTM PVC, but all analyses were repeated using BP_ND_ without PVC.

## Results

Of the fifteen patients with CBS, six had a cortical [_11_C]PiB SUVR more than 1.21 and were therefore excluded from further analysis in this paper. The remaining groups (9 CBD, 14 PSP, 15 controls) were matched in age, sex and education (Table 1). We observed typical cognitive profiles, as summarised in Table 1: patients were impaired on memory, verbal fluency, language and visuospatial domains of the ACE-R, MMSE and MoCA. There were high endorsements on the Cambridge Behavioural Inventory, and the Clinical Dementia Rating scale, with impairment of activities of daily living on the Schwab and England scale. Concurrent medications used by our participants at the time of the [_11_C]UCB-J PET scan are outlined in Supplementary Table 1. Four of our patients (1 PSP, 3 CBD) were on dopaminergic medication, and 9 on amantadine (3 PSP, 6 CBD).

Compared to controls, in patients there was a significant global reduction in [_11_C]UCB-J BP_ND_ (Figure 1A-C) across all major cortical and subcortical areas (p<0.05 FDR corrected for all regions of interest shown in Figure 1C); regional BP_ND_ values for the three groups are reported in Table 2. BP_ND_ in PSP and CBD was 20-50% lower than controls (p<0.01), with the most severe median reduction seen in the medulla, substantia nigra, pallidum, midbrain, pons and caudate nucleus in patients with PSP, and in the medulla, hippocampus, amygdala, caudate nucleus, insula and thalamus in patients with CBD. Post-hoc analysis revealed that the significant differences in BP_ND_ between patients and controls in the pallidum and substantia nigra were mainly driven by the PSP cohort. Using data without GTM PVC, the pattern of statistically significant differences in BP_ND_ for the reported regions in Table 2 remains, p<0.001.

**Figure 1:**
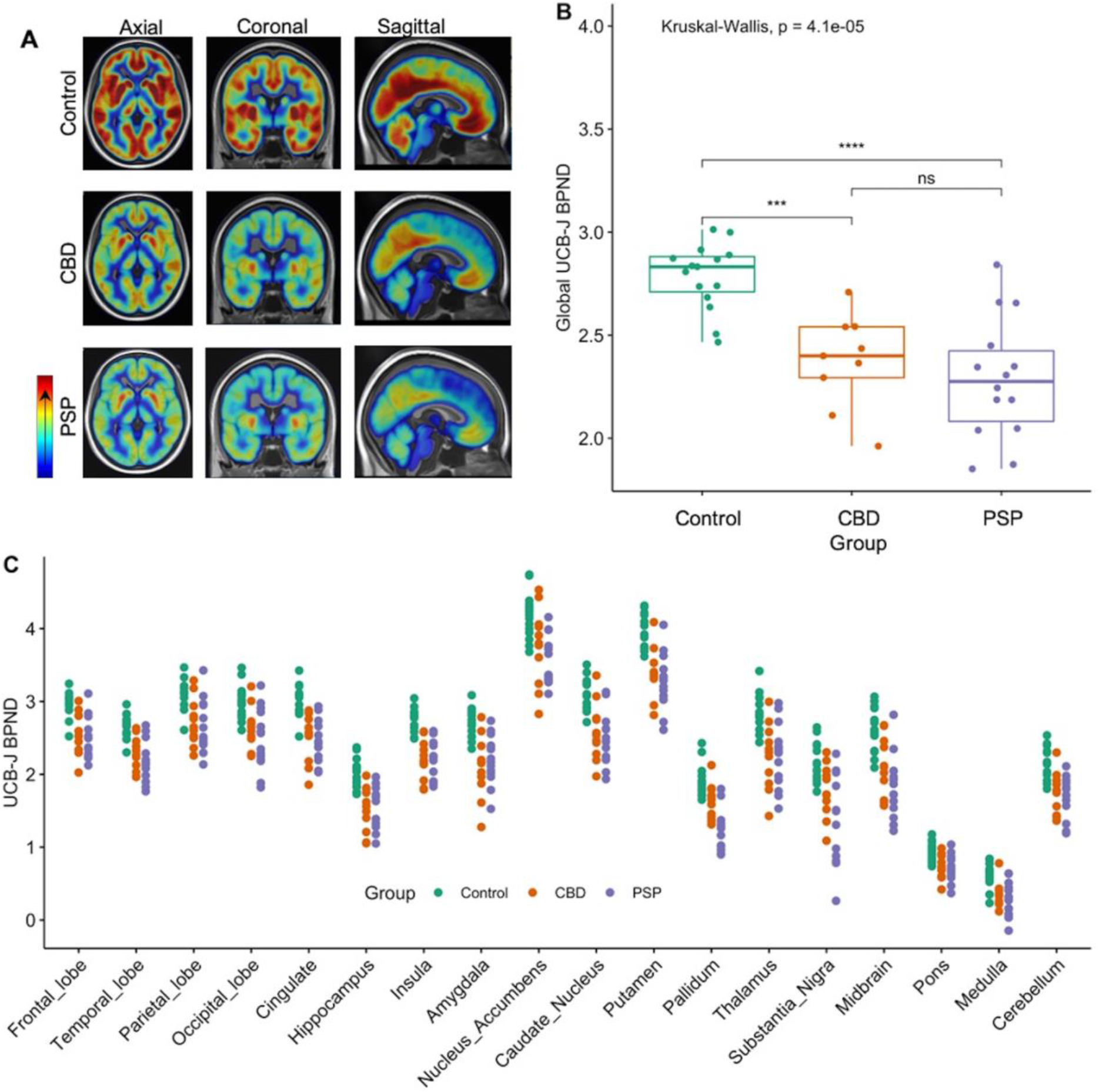
**(A)** Mean [_11_C]UCB-J BP_ND_ maps for control participants (top row), CBD (middle row), and PSP (bottom row); high and low BP_ND_ values are shown by red and blue areas, respectively. **(B)** Reduction in global [_11_C]UCB-J BP_ND_ across patients compared to controls (P<0.05). **(C)** Individual regional GTM PVC [_11_C]UCB-J BP_ND_ values for control, CBD and PSP participants, across major regions of interests. Binding potential values for patients differed significantly from controls in all the regions depicted (P<0.05, FDR corrected).

**Table 2.** Mean (standard deviation) GTM PVC [_11_C]UCB-J BP_ND_ values per group for cortical and subcortical regions of interest. (surviving false discovery rate correction over 18 regions).

| Region | Control | CBD | PSP | F (p) |
| --- | --- | --- | --- | --- |
| Frontal Lobe | 2.96 (0.17) | 2.60 (0.29) | 2.48 (0.28) | 15.05 (<0.0001) |
| Temporal Lobe | 2.68 (0.16) | 2.30 (0.23) | 2.17 (0.27) | 19.34 (<0.0001) |
| Parietal Lobe | 3.11 (0.19) | 2.75 (0.32) | 2.63 (0.36) | 10.10 (<0.0003) |
| Occipital Lobe | 2.98 (0.23) | 2.66 (0.29) | 2.48 (0.41) | 8.80 (0.0008) |
| Cingulate | 3.02 (0.21) | 2.56 (0.26) | 2.46 (0.28) | 20.41 (<0.0001) |
| Insula | 2.76 (0.15) | 2.24 (0.26) | 2.17 (0.27) | 28.55 (<0.0001) |
| Amygdala | 2.71 (0.20) | 2.18 (0.34) | 2.20 (0.33) | 14.67 (<0.0001) |
| Nucleus Accumbens | 4.18 (0.31) | 3.85 (0.46) | 3.54 (0.33) | 11.28 (0.0002) |
| Hippocampus | 2.00 (0.20) | 1.57 (0.29) | 1.57 (0.30) | 12.37 (<0.0001) |
| Caudate Nucleus | 3.12 (0.22) | 2.59 (0.41) | 2.48 (0.36) | 15.59 (<0.0001) |
| Pallidum | 1.90 (0.22) | 1.65 (0.24) | 1.27 (0.31) | 20.69 (<0.0001)<sub>a</sub> |
| Putamen | 3.99 (0.24) | 3.43 (0.32) | 3.28 (0.37) | 19.94 (<0.0001) |
| Thalamus | 2.86 (0.25) | 2.29 (0.45) | 2.25 (0.44) | 11.23 (<0.0002) |
| Cerebellum | 2.13 (0.22) | 1.75 (0.30) | 1.69 (0.28) | 11.50 (0.0001) |
| Midbrain | 2.61 (0.29) | 2.16 (0.38) | 1.83 (0.42) | 16.61 (<0.0001) |
| Substantia Nigra | 2.13 (0.28) | 1.72 (0.34) | 1.32 (0.59) | 12.79 (<0.0001)<sub>a</sub> |
| Pons | 0.93 (0.13) | 0.75 (0.18) | 0.71 (0.18) | 7.69 (0.002) |
| Medulla | 0.62 (0.16) | 0.37 (0.17) | 0.28 (0.24) | 12.04 (<0.001) |

CBD here refers to corticobasal syndrome with a negative amyloid biomarker from [ $^{11}\text{C}$ ]PiB PET, and PSP refers to patients with PSP-Richardson's syndrome. F-statistic and p-values derived from an ANCOVA across the three groups, with age as a covariate of no interest. The significant difference here is driven by the PSP group only.

The reduction in synaptic density was seen even in the areas of the brain that did not show significant grey matter atrophy. Figure 2A shows the group differences in grey matter volume normalized against the mean of the control group; the significant areas of grey matter volume loss were in the caudate nucleus (p=0.01), and thalamus (p=0.04) in the CBD cohort, and in the frontal (p<0.01), temporal (p=0.04), parietal (p<0.01), occipital lobes (p<0.01), caudate nucleus (p<0.001), and the thalamus (p<0.01) in the PSP cohort. The reduction in [_11_C]UCB-J BP_ND_ however, is more extensive and consistently significantly different across all major cortical and subcortical areas as shown in the normalised plot in Figure 2B (binding potentials were normalised against the mean binding potential of the control cohort for each region of interest).

**Figure 2:**
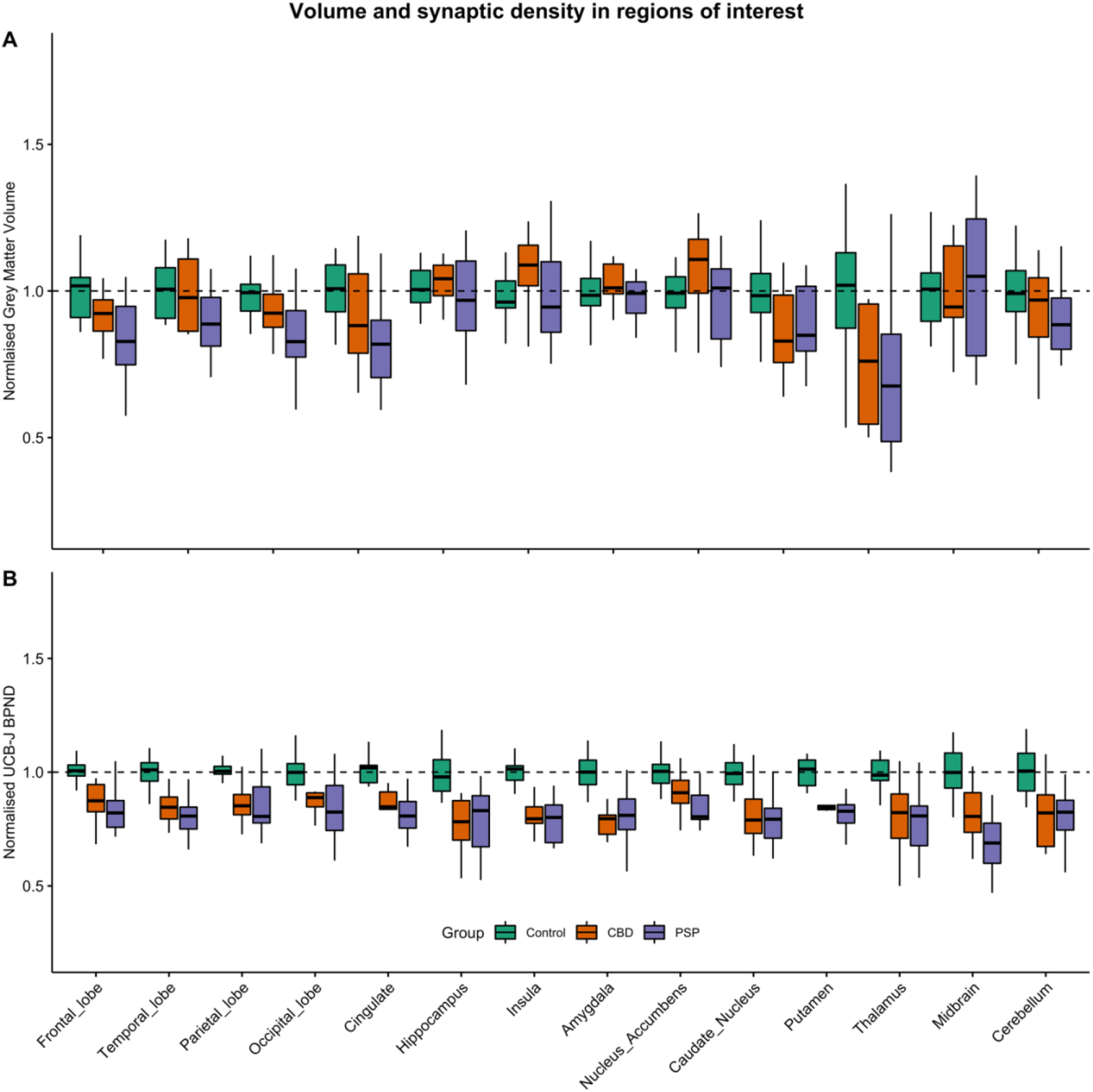
**(A)** Cortical and subcortical grey matter volumes, normalised against the corresponding volumes in controls, were significantly reduced in the caudate nucleus and thalamus in CBD; and in frontal, temporal, parietal and occipital lobes, as well as in the caudate nucleus, and thalamus in PSP, P<0.05. **(B)** Mean-centred [_11_C]UCB-J BP_ND_ across cortical and subcortical regions of interest normalised against the corresponding BP_ND_ values in controls, demonstrating a median reduction of 20-50%.

Correlations between [_11_C]UCB-J BP_ND_ and both global cognition and disease severity are given in Figure 3. A significant positive correlation was seen between [_11_C]UCB-J BP_ND_ and the revised Addenbrooke’s Cognitive Examination total score (R=0.52, p=0.01) (Figure 3A). There was a significant negative correlation between [_11_C]UCB-J binding and the PSP (R= − 0.61, p<0.01), and CBD (R= −0.72, p<0.001) rating scales (Figures 3B and 3C)

**Figure 3:**
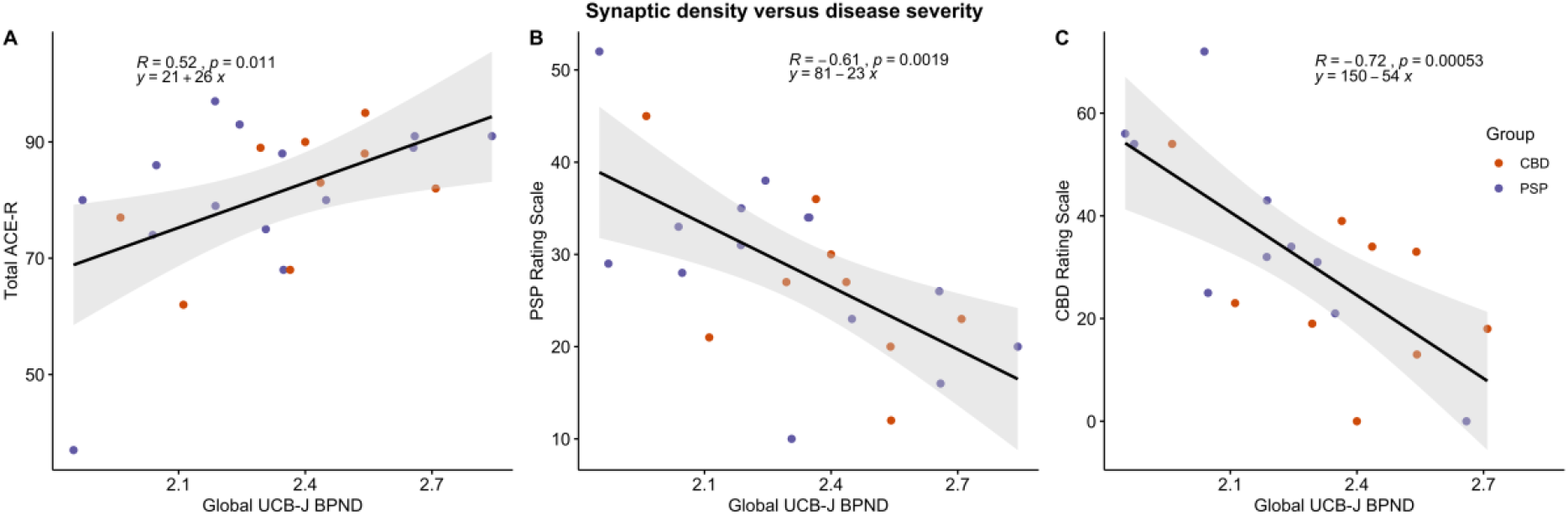
Correlations between global [_11_C]UCB-J BP_ND_ and total ACE-R score (A), total PSP rating scale (B), and total CBD rating scale (C) for the two patient groups.

## Discussion

The principal result of this study is a widespread reduction in synaptic density in PSP-Richardson’s syndrome and amyloid-negative Corticobasal Syndrome (CBS) (which we define as Corticobasal Degeneration (CBD). This accords with *post mortem* estimates of synaptic loss in PSP and CBD, using synaptophysin immunohistochemistry (6), imaging of neurite density in PSP (41) and morphological studies of cortical dendrites in the closely related condition of frontotemporal lobe dementia (42). Indirect evidence of synaptic loss, from consequential reduction in metabolism, comes from [_18_F]FDG PET changes in frontal, temporal and parietal lobes (43-46). However, PET imaging with the ligand [_11_C]UCB-J provides direct evidence *in vivo* of severe and extensive loss of cortical and subcortical synapses, including areas of the brain that are minimally atrophic (47).

Progressive supranuclear palsy and corticobasal degeneration are progressive, with an average disease duration of five to eight years from symptom onset (48). In our clinically diagnosed CBD and PSP groups, the mean symptom duration at the time of PET was three and a half years, and our patients were likely to be approximately mid-way through their symptomatic disease course (not including a potentially long pre-symptomatic period). The median reduction of 20% (and maximal 50%) in [_11_C]UCB-J binding observed *in vivo* compared to controls, is therefore in keeping with the predictions from *post mortem* data.

The synaptic loss observed in our study is widespread, extending beyond the regions that are arguably most associated with the diseases. In PSP, from *post mortem* studies, these include basal ganglia, thalamus, substantia nigra, premotor cortex, as well as the dentate nucleus and cerebellar white matter. In CBD, areas associated with the disease include cortex, thalamus, basal ganglia and brainstem, without cerebellar involvement (48-50). However, in our study the loss of synapses in PSP is global across the cortex, and not confined to the premotor and motor areas, and extends beyond the substantia nigra in the brainstem with pontine and medullary involvement. The loss of synapses in the cerebellum in PSP echoes pathological studies of tau distribution in this disease (49). Interestingly, the cerebellum was also markedly abnormal in CBD; although cerebellar atrophy and tau accumulation are not typical associations of CBD (49). Cerebellar synaptic loss in CBD may therefore represent cerebellar diaschisis in response to widespread cortical pathology and loss of cortico-cerebellar projections; a small minority of individuals in an amyloid-negative CBS cohort may have PSP as the underlying cause for their corticobasal syndrome, although this is unlikely to be sufficient to drive the group-wise effect.

Preclinical models of tauopathy suggest early synaptotoxicity with reduced plasticity and density (ii), in response to soluble oligomeric tau aggregates (12) and inflammation (51). The toxicity associated with tau pathology leading to synapse loss is complex and involves direct and indirect pathways (reviewed in Spires-Jones *et al. 2014* (52)). Naturally occurring tau plays a role in synaptic function through modulating microtubule and axonal stability; disruptions to this machinery leads to prevention of the trafficking of essential components to synapses such as synaptic receptors (53) and mitochondria. Indeed, over-expression of tau interferes with mitochondria transport (54), and contributes to hyperexcitability of neurons and impaired calcium influx in transgenic mouse models (rTg4510) (55). The global nature of synaptic reduction suggests a more widespread pathology in the primary tauopathies of PSP and CBD beyond the areas that are histologically reported as harbouring a high tau burden such as the basal ganglia, thalamus, and brain stem (56). This may in part be explained by the global damage caused by oligomers of tau which are not easily visible on tau PET imaging or histology. In support of this are biochemical studies that report tau accumulation in both grey and white matter by western blot in PSP but not necessarily by immunohistochemistry (57).

We observed a significant correlation between synaptic loss and disease severity in PSP and amyloid-negative CBS. Synaptic loss correlates with cognitive impairment in another clinical tauopathy, Alzheimer’s disease (13,58), and preclinical models of this (59,60). Our *in vivo* PET results support the potential use of synaptic PET as a marker of disease and progression, but longitudinal data are required. Synaptic PET may support early stage clinical trials in PSP and CBS/CBD; it is encouraging in this latter respect that [_11_C]UCB-J PET is sensitive to changes in synaptic density, for example in response to treatment with the synaptic modulator Saracatinib (61).

Our study has several limitations. Although the sample size is small, it is adequately powered in view of the large effect sizes predicted. However, subtler relationships with mild disease, progression or individual clinical features, or phenotypic variants of PSP and CBS, require larger studies. We acknowledge the potential for off-target binding, but preclinical data indicate very high correlations between UCB-J and synaptophysin, a marker of pre-synaptic vesicular density (25). Our diagnoses were clinical, without neuropathology, although the clinicopathological correlations of PSP-Richardson syndrome are very high, and in the absence of Alzheimer’s disease, the clinicopathological correlation of CBS with a 4R-tauopathy (CBD or PSP) is also high (18). Binding potentials for SV2A radioligands such as [_11_C]UCB-J can be confounded by the use of concurrent medication that may bind to SV2A. We did not enrol any individuals taking levetiracetam or any member of this family of drugs that are SV2A-specific ligands (62). Previously reported studies using [_11_C]UCB-J in disease have usually not commented on medications used by participants, however one study using this ligand in major depressive disorders reports exclusion of participants on psychotropic medications in the 2 months preceding PET scanning (63); whilst many of our PSP and CBD patients are on medications falling under the psychotropic umbrella, to our knowledge, none of these bind to SV2A.

Arterial blood sampling was not carried out in this study; we used reference tissue modelling to reduce the demand on our patient cohort. Reference tissue modelling of [_11_C]UCB-J with the centrum semiovale as the reference tissue has been verified against arterial input function compartmental modelling in healthy controls (39,40) and in Alzheimer’s disease (64). To assess the validity of the centrum semiovale in our cohort, we determined the mean total distribution volume (V_T_) for each of our subject groups using standard arterial input function data from the literature (26,65); this approach assumed that the standard input function was equally valid for all groups. This analysis indicated a small positive bias in centrum semiovale V_T_ for CBD (5%) and less so PSP (2%) relative to that in controls, which would lead to a commensurate reduction in BP_ND_ under the assumption that the non-displaceable distribution volume (Vnd) in the target ROIs remains invariant. These biases cannot, however, explain the much greater BP_ND_ reductions seen for CBD and PSP, which is especially true for PSP. Indeed, scaling BP_ND_ in the CBD and PSP cohorts to account for the above biases in centrum semiovale V_T_, produced a similar pattern of significant global reduction in BP_ND_ for patients compared to controls, except that the significant differences in the midbrain, pons, substantia nigra, pallidum, and occipital lobe were primarily driven by the PSP cohort in the post-hoc analysis.

The therapeutic challenge in tauopathies is partly due to the complex nature of the underlying pathology. Early stage trials will require early accurate diagnosis, although diagnosis is typically made 3 years after symptom onset (66,67). It is unlikely that synaptic PET could provide pre-symptomatic diagnosis in rare conditions, but it is a promising tool to characterise pathogenetic mechanisms, monitor progression and assess response to experimental medicines (68).

## Data Availability

The derived data that support the findings of this study are available from the corresponding author, upon reasonable request for academic (non-commercial) purposes.

## Acknowledgements

The authors would like to thank the participants, the staff at the Wolfson Brain Imaging Centre, and the staff at the Cambridge Centre for Parkinson-Plus. We thank the NIHR Cambridge Biomedical Research Centre for Support.

## Authors’ Roles

1. Research project: A. Conception, B. Organization, C. Execution;
2. Statistical Analysis: A. Design, B. Execution, C. Review and Critique;
3. Manuscript: A. Writing of the first draft, B. Review and Critique.

Negin Holland (1A-C, 2A-C, 3A-B); P. Simon Jones (2C, 3B); George Savulich (1B-C, 3B); Julie K. Wiggins(1B-C,3B); Young T. Hong (1C, 2C, 3B); Tim D. Fryer (2C, 3B); Roido Manavaki (1C, 2C); Selena Milicevic-Sephton (1C, 3B); Istvan Boros (3C); Maura Malpetti (2C, 3B); Frank H. Hezemans (2B, 3B); Franklin I. Aigbirhio (1A, 3B); Jonathan P. Coles (1A, 3B); John O’Brien (1A, 2C, 3B); James B. Rowe (1A, 2C, 3B).

## Financial Disclosures of all authors (for the preceding 12 months)

NH is funded by the Association of British Neurologists - Patrick Berthoud Charitable Trust. PSJ has no financial disclosures to report. GS has no financial disclosures to report. JKW has no financial disclosures to report. YTH has no financial disclosures to report. TDF has no financial disclosures to report. RM has no financial disclosures to report. SMS has no financial disclosures to report. IB has no financial disclosures to report. FIA has no financial disclosures to report. JBR serves as an associate editor to Brain, and is a non-remunerated trustee of the Guarantors of Brain and the PSP Association (UK). He provides consultancy to Asceneuron, Biogen and UCB and has research grants from AZ-Medimmune, Janssen and Lilly as industry partners in the Dementias Platform UK. JOB provides consultancy to Axon, TauRx and Eisai and has research grant support from Alliance Medical and Merck.

## Notes

**Financial Disclosures/conflict of interest:s** The authors do not have any competing interest pertaining to the manuscript.

**Funding:** The study was funded by the Cambridge University Centre for Parkinson-Plus; the National Institute for Health Research Cambridge Biomedical Research Centre (SUAG/004 RG91365 JBR); the Wellcome Trust (103838) and the Association of British Neurologists, Patrick Berthoud Charitable Trust (RG99368). MP is supported by Cambridge Trust Vice-Chancellor’s Award & Sidney Sussex College Scholarship. FHH is supported by a Cambridge Trust Vice-Chancellor’s Award & Fitzwilliam College scholarship.

### Funding Statement

The study was funded by the Cambridge University Centre for Parkinson-Plus; the National Institute for Health Research Cambridge Biomedical Research Centre (SUAG/004 RG91365 JBR); the Wellcome Trust (103838) and the Association of British Neurologists, Patrick Berthoud Charitable Trust (RG99368).

